# Beyond legalization: characterizing distinct recreational cannabis regulatory approaches across US states, 2013-2024

**DOI:** 10.64898/2026.02.26.26346986

**Authors:** Ariadne Rivera-Aguirre, Ellicott C. Matthay, Álvaro Castillo-Carniglia, Silvia S. Martins, Iván Díaz, Magdalena Cerdá

## Abstract

**Background:** Recreational cannabis legalization has expanded rapidly across US states. The regulatory approaches states adopt vary widely, with varying implications for public health. This study aimed to characterize heterogeneity in recreational cannabis laws (RCLs) across US states and to identify state-level characteristics associated with these regulatory models.

**Methods:** We conducted Latent Class Analysis (LCA) of state-year RCL provisions from 2013 to 2024 (n=612) to identify distinct RCL approaches. Descriptive analyses and exploratory multinomial regression analyses were used to examine correlations between state characteristics and RCL approaches from 2020 to 2024, when sufficient cross-state variation in RCL adoption was available.

Eleven recreational cannabis policy provisions spanning governance, potency limits, consumption restrictions, access controls, taxation, marketing regulations, and driving prohibitions are primarily from the Alcohol Policy Information System. State-level characteristics included cannabis use prevalence, market conditions, medical cannabis history, political factors, demographic, and socioeconomic covariates obtained from multiple secondary data sources.

**Results:** We identified four latent classes of state-year RCL provisions representing different regulatory approaches: No RCL, Pre-commercial, Full Access, and Dispensary Access. The No RCL corresponded to state-years without RCL. The Pre-commercial class represented state-years in early-stage legalization with a minimal regulated approach in terms of commercial infrastructure. The Full Access class was characterized by permitting on-site retail consumption and home delivery and restricting (but not prohibiting) public use. In contrast, the Dispensary Access class limited retail sales to off-site consumption only, prohibited public use, and imposed stricter market controls.

Higher past-month cannabis use prevalence was associated with a greater likelihood of membership in the Full Access class (RRR = 1.78; 95% CI: 1.21–2.62), relative to No RCL. A longer duration since medical cannabis legalization was associated with a higher likelihood of membership in the Dispensary access class (RRR = 1.47; 95% CI: 1.02–2.12). Higher beer excise taxes were associated with a lower likelihood of membership in any RCL class relative to No RCL.

**Conclusions:** From 2013 to 2024, US recreational cannabis regulations clustered into four distinct regulatory approaches, with two distinct commercial models: one permitting on-site retail consumption and home delivery, the other restricting sales to off-premises only and prohibiting public use. Higher cannabis use prevalence and longer medical cannabis history were associated with more access-oriented and more restrictive commercial approaches, respectively.

## 1. BACKGROUND

Over the past two decades, the regulatory landscape of cannabis for recreational use in the US has been rapidly changing. In 2012, Colorado and Washington became the first two states to legalize the use of cannabis for recreational purposes. Since then, 22 additional states and the District of Columbia have followed (1). As cannabis shifts from an illicit substance to a regulated substance, the effects of the rules governing the use and accessibility of recreational cannabis have become an area of public health concern. The effect of the new policies regulating recreational cannabis on public health will likely depend on the specific provisions that jurisdictions adopt (2–4). While some regulatory approaches might increase adverse effects associated with cannabis use, such as cannabis use disorder, other policy choices, including potency limits and potency-based taxes, may be more successful in mitigating the negative impacts associated with cannabis use (5). Thus, research is needed to understand the variation in the recreational policy provisions that states are adopting.

As of November 2025, most US states that have legalized cannabis for recreational use have adopted a commercial model of recreational cannabis legalization, characterized by licensed private production and retail sales (except for Washington, DC) (1). However, the specific provisions within recreational cannabis laws (RCLs) have been far from uniform (6,7). States have implemented different approaches to regulating cannabis production and supply of cannabis products, including taxation schemes, pricing and marketing restrictions, and the types of products that may be sold (7,8). The characteristics of these policies have also evolved (5,6,9). The early regulatory approaches considered a few general aspects aligned with public health objectives, such as preventing the distribution of cannabis to minors and preventing impaired driving (5,10). However, over time, regulators have increasingly adopted additional public health-related policy provisions, including warning-labeling requirements, marketing restrictions, and taxes based on THC concentrations in products sold (5).

Despite the growing number of states legalizing recreational cannabis use, few studies have characterized the variation in the nature of RCL policy provisions across states and time (6). Research on policy heterogeneity in the cannabis landscape has primarily focused on Medical Cannabis Laws (MCLs) (11,12). A comprehensive understanding of what states have adopted varying marketing and advertising restrictions, retail access rules, and quantity and potency limits in RCL contexts—and how these patterns have evolved—is still needed (13–17). To our knowledge, two studies have assessed heterogeneity in US state cannabis policy environments, including RCLs, using a scoring system (6,18). Although composite scores allow identification of overall policy restrictiveness, the additive nature of the scores may obscure which specific policies drive those differences.

Additionally, the policy provisions have not been adopted by states; they tend not to occur randomly. Prior studies have noted that states with larger commercial markets tended to legalize recreational cannabis through ballot initiatives (9). The passage of RCLs may be consistent with policy learning and diffusion theories in several ways (19,20). First, states may regulate recreational cannabis by adopting regulatory approaches similar to those for other substances, such as tobacco and alcohol (21,22). Second, as more adjacent and peer states adopt specific recreational cannabis provisions, the likelihood that a state will adopt those same provisions may increase (23). Third, states with established medical programs are more likely to transition to RCLs (3). Social, economic, and political characteristics, such as political affiliation, age composition, and income, may also drive differences in specific RCL provisions across states and over time (3,24–26). Examining the relationship between state-level characteristics and RCL types would help identify the structural and contextual drivers of policy differences and their implications for public health in emerging cannabis markets (23,27–29).

In this study, we aimed to advance understanding of RCLs by developing a typology of recreational cannabis policies from 2013 to 2024, with a specific focus on demand-side, consumer-facing regulations most proximal to public health outcomes (2,27,30,31). Using latent class analysis (LCA), we employed a data-driven approach to classify state-years into latent policy regimes that capture common combinations of consumption restrictions, potency limits, pricing controls, and other provisions likely to be adopted concurrently. By extending the measures through 2024, we provided a contemporary characterization of the typologies of RCLs adopted across US states over time. Additionally, we explored the relationship between state-level characteristics and distinct recreational cannabis regulatory approaches to further the understanding of the factors correlated with specific regulatory approaches during the later period of broader RCL adoption (2020–2024).

## 2. Methods

### 2.1 Data

#### Recreational cannabis policy characteristics

We analyzed annual recreational cannabis policies from January 1^st^, 2013, to January 1^st^, 2024. The primary data source was the Alcohol Policy Information System (APIS) from the National Institute of Alcohol Abuse and Alcoholism (NIAAA), which provides state-level legal data on recreational cannabis policies. We obtained adoption dates of laws permitting home cultivation of recreational cannabis from Martins et al. (32). Policies were assigned to a specific year if they were in effect as of January 1^st^ of that year. We cross-referenced the dataset with data from the Network for Public Health Law (33), Marijuana Policy Project (34), and state websites.

We considered 16 RCL provisions that captured core dimensions of recreational cannabis legalization guided primarily by the National Academies of Sciences, Engineering, and Medicine’s (NASEM) (5) Report on “Cannabis Policy Impacts Public Health and Health Equity”, which identified a set of best practices likely to be protective of public health. We complemented these policy items with additional policy dimensions identified in the health policy literature as protective approaches to health (9,10,13,37,38). We selected 16 RCL provisions spanning seven regulatory domains (Table 1): governance, permitted products, access and retail controls, consumption restrictions, pricing and taxation, marketing, youth restrictions, and driving restrictions. The governance domain captured whether regulatory agencies included health or consumer protection departments, and the role of local authorities in implementation. The products permitted domain referred to jurisdictions allowing retail sales of concentrates or edibles, and whether any THC limits were set for retail products. Access and retail control encompassed provisions on home cultivation, home delivery, and statewide limits to the number of retail establishments. Consumption restrictions referred to public-use restrictions, and whether the jurisdiction allowed sales for off-premises consumption only, or sales for both on- and off-premises consumption. Pricing and taxation provisions included pricing controls and the structure of taxation regimes. Marketing provisions captured requirements for packaging, labeling, and advertising. Youth restrictions referred to prohibitions on furnishing, possession, consumption, and purchase by persons under 21 years of age. Lastly, the driving restrictions domain covered adult impaired-driving prohibitions and open-container regulations in passenger compartments. We excluded supply-side policies available in APIS related to commercial license cultivation restrictions, vertical integration bans, and seed-to-sale tracking requirements. While these measures may indirectly influence pricing and availability, they are more distantly related to consumption behaviors and thus public health. Excluding these measures allowed us to maintain a conceptual focus on provisions more directly linked to demand-side regulations and to avoid over-parametrizing our models relative to the sample size. Table S1 in the Appendix lists the exact provisions and the operationalizations used in this study.

**Table 1.** Recreational cannabis policy domains and provisions.

| Domain | Provision |
| --- | --- |
| Governance | Regulatory agency<br>Role of local authority<sup>1</sup> |
| Products permitted | THC limits<sup>1</sup> |
| Access and retail control | Home cultivation<br>Home delivery<sup>1</sup><br>State-wide retail outlet restrictions |
| Consumption restrictions | Public use restrictions<sup>1</sup><br>Permitted consumption setting for retail cannabis<sup>1</sup> |
| Pricing and Taxation | Pricing controls<sup>1</sup><br>Taxation regimes<sup>1</sup> |
| Marketing | Packaging requirements<sup>1,2</sup><br>Warning labeling requirements<sup>1,2</sup><br>Advertising restrictions<sup>1</sup> |
| Underage restrictions | Underage restrictions |
| Driving restrictions | Adult impaired driving prohibitions<sup>1</sup><br>Open containers in passenger compartments<sup>1</sup> |

<sup>1</sup>Provision included in the final LCA model
<sup>2</sup>Packaging and warning labeling provisions were combined prior to analyses

#### State characteristics

To investigate state-level characteristics associated with the adoption of specific RCL approaches, we compiled annual state-level measures from 2019 to 2023. Although RCL data were available from 2013, we restricted the analysis to the later period, as RCL was more widely adopted, thereby ensuring meaningful variation across RCL approaches. We hypothesized that the types of RCLs that states adopted between 2020 and 2024 would be correlated with distinct market conditions, prior cannabis policy measures, the broader regulatory environment for other substances, political factors, and the state’s demographic and socioeconomic composition.

Previous studies have found that states with legalized cannabis had higher baseline levels of cannabis use (39). To determine whether pre-existing market conditions were associated with the type of RCL adopted, we analyzed the correlation between past-month cannabis use among individuals aged 12 years and older and the categories of RCLs adopted across states (40,41).

Building on the concept of policy diffusion, the process of learning, imitation, and competition among jurisdictions may have also influenced the types of RCLs that states adopted (19,20,42). We thus examined associations between RCL types and cannabis policy diffusion, including the proportion of neighboring states with any RCLs (33), cannabis sales per capita in the state, the number of years since medical cannabis legalization (35), and whether medical cannabis outlets were operational in the state (35).

To capture broader state approaches to substance regulation, we included the state excise tax rate on beer (43) and the presence of comprehensive smoke-free laws (44). Prior studies have found that direct-democracy mechanisms and political context predict recreational marijuana adoption (23). Therefore, we also considered other policy and political factors, including the percentage of votes in the 2016 and 2020 presidential elections in favor of the Republican Party (45), the density of social organizations (e.g., religious organizations, charities, interest groups) (46), the adoption of Medicaid expansion (47), whether the state allowed for citizen-initiated ballots (48), and the revenue-to-expenditure ratio as a measure of state financial health (49).

Demographic and socioeconomic factors may also shape the political feasibility of and demand for RCLs. The demographic factors we considered were the percentage of the state population that is non-Hispanic White, the percentage of the state population that is male, the median age, and the percentage of the population residing in an urban area (46). The socioeconomic characteristics we considered were the percentage of the state population aged 25 and older with at least a high school education, median household income, the Gini coefficient of income inequality, and the unemployment rate (46,50). A comprehensive list of variables, operationalization, and data sources is presented in Table S2 in the Appendix.

### 2.2 Analytic approach

We implemented a standard latent class analysis (LCA) (51) to identify state-year subgroups (“latent classes”) of recreational cannabis regulatory approaches based on the policy provisions in Tables 1 and S1. The standard LCA approach treats each state-year as an independent observation, maximizing statistical power to detect policy patterns. While MLCA accounts for within-state correlation, it requires larger upper-level sample sizes and additional parametrization that would limit power and model stability (52) given our 51-state sample size. We implemented MLCA as a sensitivity analysis (described below).

We applied an iterative process, removing provisions that showed no between-class differentiation and re-estimated the model until all provisions contributed meaningfully to class separation. The final LCA was composed of 11 cannabis policy provisions: role of local governments, potency limits, consumption restrictions, access, taxation and pricing controls, marketing, and driving prohibitions. We fit models with 1 to 5 classes. Following best practices, model fit was evaluated using multiple fit indices: Bayesian Information Criterion (BIC), sample size adjusted BIC (aBIC), Consistent Akaike Information Criterion (CAIC), Vuong-Lo-Mendell-Rubin (VLMR) test, and approximate correct model probability (cmPk) (53,54).Since simulation studies indicate that no single fit measure identifies the correct solution, we used this comprehensive set of metrics. In cases of disagreement, we prioritized the solution where BIC minimized, VLMR remained significant, and classes were substantively interpretable (53,54).

After identifying the optimal number of classes, we first described the distribution of state-level characteristics lagged by 1 year across the latent classes from 2020 to 2024. Subsequently, we estimated multinomial logistic regression with clustered standard errors to assess the associations between state characteristics and latent policy classes. Each state-year was assigned to its most likely latent class based on posterior probabilities. Although no minimum probability threshold was imposed, posterior classification probabilities averaged >0.95 across classes, indicating a high degree of classification certainty. We excluded state characteristics with low variation or quasi-complete separation (e.g., one or more latent policy classes had few or no observations for a given covariate category). We examined covariate redundancy and multicollinearity diagnostics. High variance inflation factor (VIF>10) and pairwise correlations (>0.8) were used to guide the selection of the most theoretically relevant predictors to resolve the convergence issues. The characteristics considered but not included due to multicollinearity, sparsity, or low variance were whether medical cannabis outlets were operational, smoke-free laws, Medicaid expansion, whether citizen ballot initiatives were allowed, state finance measures, the percentage of the population that was male, and the percentage of the population with a High School diploma or more. Lastly, we also considered the number of retail cannabis outlets. However, since we did not have data available for all states in our study, we excluded this measure from the correlation analysis to preserve sample size. Still, we included it in the descriptive results Table 2.

**Table 2.** State-year descriptive characteristics (2019–2023) by recreational cannabis regulatory approaches, 2020-2024.

| Characteristic | no RCL | Pre-commercial Model | Full Access Model | Dispensary Access |
| --- | --- | --- | --- | --- |
| *Cannabis market* |  |  |  |  |
| Past-month cannabis use (%) (Ages 12+) | 9.60 (2.18) | 14.23 (4.23) | 15.51 (3.20) | 16.08 (3.48) |
| Annual cannabis sales per capita (\$) | 22.01 (45.31) | 24.33 (26.03) | 177.25 (144.45) | 138.42 (86.66) |
| Dispensaries per 10,000 people* | 0.12 (0.44) | 0.17 (0.42) | 0.59 (0.56) | 0.51 (0.57) |
| *Cannabis policy adoption and diffusion* |  |  |  |  |
| Neighboring states with RCL (%) | 58.3 (49.3) | 68.8 (46.4) | 58.8 (49.2) | 71.4 (45.2) |
| Years since MCL | 2 [0–6.50] | 11 [7.25–13] | 15 [11.25–22] | 20.5 [13.25–24] |
| Medical cannabis outlets operational (%) | 49.1 (50) | 81.2 (39) | 79.4 (40.4) | 100 (0) |
| *Substance use and health policy* |  |  |  |  |
| Beer excise tax (USD per gallon) | 0.35 (0.32) | 0.17 (0.09) | 0.29 (0.34) | 0.26 (0.20) |
| Smoke-free laws in effect (%) | 63.2 (48.2) | 75.0 (43.3) | 85.3 (35.4) | 97.6 (15.2) |
| State adopted Medicaid expansion (%) | 57.1 (49.5) | 100 (0) | 100 (0) | 100 (0) |
| *Political characteristics* |  |  |  |  |
| Citizen ballot initiatives allowed (%) | 30.1 (45.9) | 56.2 (49.6) | 52.9 (49.9) | 69 (46.2) |
| Votes in favor of the republican party in presidential elections (%) | 53.87 (9.43) | 32.19 (20.29) | 42.90 (6.68) | 41.28 (6.93) |
| Social organizations per 10,000 people | 10.91 (2.46) | 22.70 (17.46) | 9.32 (2.09) | 9.73 (3.04) |
| Revenue-to-expenditure ratio | 0.89 (0.11) | 0.64 (0.45) | 0.91 (0.12) | 0.87 (0.12) |
| *Demographic and socioeconomic characteristics* |  |  |  |  |
| Male population (%) | 49.52 (0.76) | 48.82 (0.93) | 49.96 (1.20) | 49.70 (0.50) |
| Median age (years) | 38.49 (2.27) | 38.23 (3.01) | 38.41 (1.75) | 39.82 (2.40) |
| Non-Hispanic white population (%) | 68.95 (15.43) | 59.69 (20.27) | 60.04 (11.91) | 67.49 (17.50) |
| Urban population (%) | 71.22 (12.06) | 79.76 (20.72) | 82.84 (10.02) | 76.05 (20.64) |
| Population 25 or older with High School+ (%) | 88.66 (2.49) | 89.13 (1.64) | 88.90 (2.30) | 88.92 (3.10) |
| Median household income (in \$1,000) | 65.78 (11.27) | 80.30 (15.29) | 79.32 (13.30) | 74.92 (10.42) |
| Unemployment rate (%) | 4.30 (1.80) | 5.01 (2.18) | 5.00 (1.90) | 4.71 (2.32) |

Values are mean (SD) for continuous variables, % (SD%) for binary variables, and median [IQR] for discrete measures.
\*Does not include data for Hawaii, Illinois, Iowa, Louisiana, Maine, Maryland, Tennessee, Vermont, Virginia, Washington, or West Virginia.

LCA model estimates were conducted using Mplus 8.11 (Muthén & Muthén, 2017) via MplusAutomation (55) in R 4.4.2 (56). The multinomial regression models were conducted in Stata 18 (57).

### 2.3 Sensitivity analysis

First, to account for the potential lack of independence among state-year units due to repeated observations within states, we conducted a sensitivity analysis using a multilevel latent class analysis (MLCA) framework. After identifying the 4-class solution from the standard LCA model presented in the main analysis, the MCLA implemented a two-level latent class model using a maximum likelihood estimator, allowing class membership probabilities to vary across states via a continuous latent factor model (58,59). Second, since DC is the only jurisdiction without recreational cannabis sales throughout the study period, we repeated analyses excluding DC. Third, to test the stability of classes over time, we repeated the standard LCA, restricting to state-years between 2020 and 2024, which corresponded to the period with the largest number of states with RCLs and a phase of greater policy maturation and differentiation, and estimated models both across all years pooled and separately by year. We also conducted an LCA separately for the subset of states that had RCLs at any point during the study period.

## 3. RESULTS

### 3.1 Classification of RCL state-years based on LCA results

Table S3 in the Appendix presents the LCA model fit for solutions ranging from 1 to 5 classes. Model fit statistics supported the 4-class LCA solution. Additionally, the 4-class solution provided a theoretically meaningful interpretation, aligning with substantive domain knowledge and distinct state-year subgroups.

Figure 1 illustrates the classification of state-years according to the 4-class solution by state and year. The 4-class model correctly identified the subgroup of state-years without RCLs Class 1 (No RCL), representing 79% of the state-years. Within state-years with RCLs, the LCA model identified three additional subgroups, each in a distinct class representing a distinct combination of regulatory provisions. Class 2, the subgroup of states with a Pre-commercial approach, constituted 5% of the sample. This subgroup typically corresponded to initial stages of legalization, before the start of retail sales. Class 3 accounted for 6% of the state-years in the study, corresponding to a Full Access approach. Class 4 accounted for 11% of the state-years in the study and represented a Dispensary Access approach. The timing of latent classes showed that states that legalized earlier were more often observed in Class 2 (Pre-commercial) and later in Class 4 (Dispensary Access approach), whereas states that legalized more recently entered directly into Class 3 (Full Access approach) or Class 4 (Dispensary Access approach).

**Figure 1.**
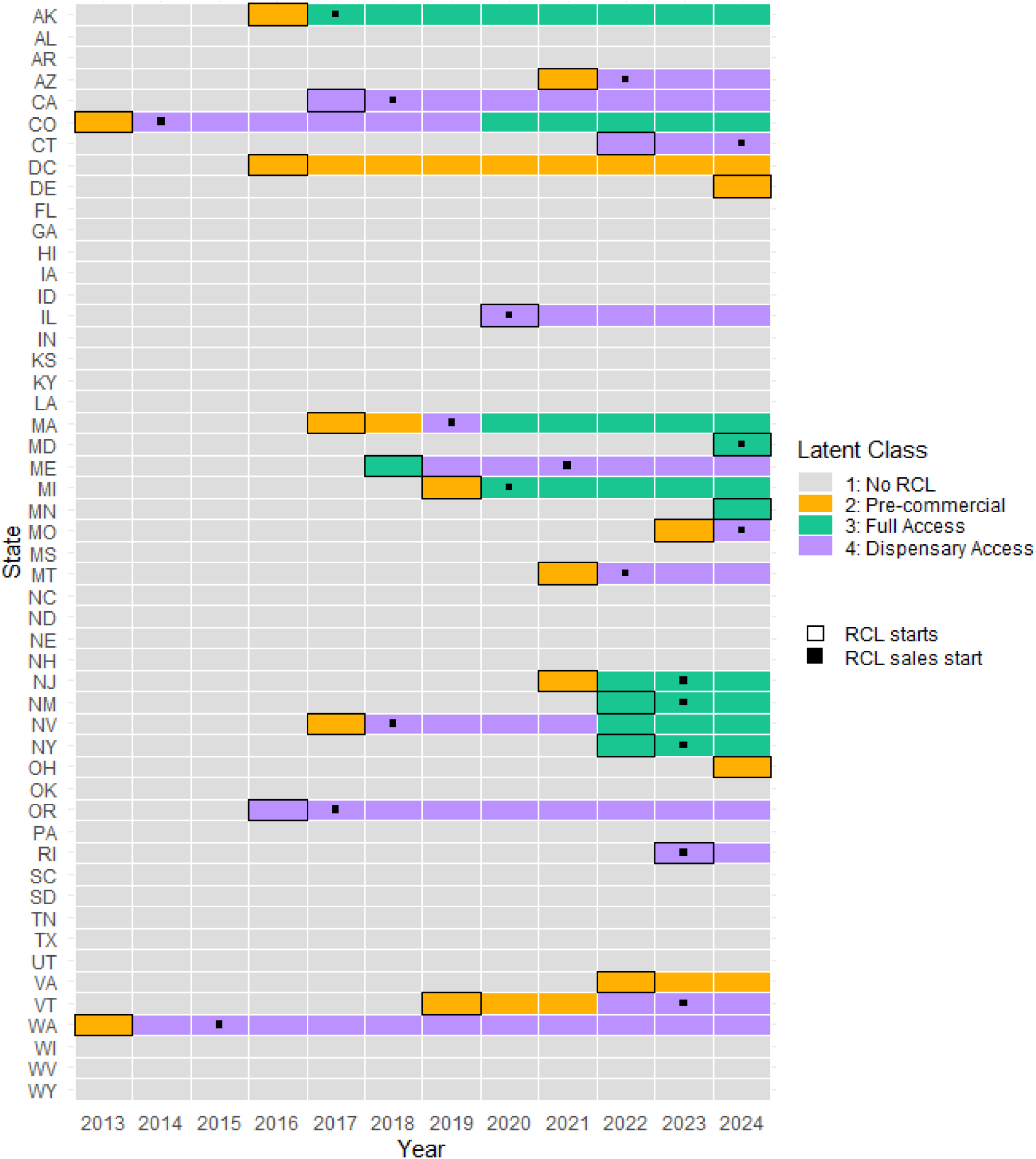
Results of the 4 latent class solution by state and year from 2013 to 2024

The item endorsements of the specific cannabis policy characteristics for each latent class are shown in Figure 2 and Table S4 in the Appendix. The No RCL class had a probability of one for absence of all provisions considered in the final model; therefore this class was excluded from Figure 2. The Pre-commercial class corresponded to state-years in which local authorities had limited regulatory rights within their borders (p=0.64), and public use was typically prohibited (p=0.8), but THC limits, advertising restrictions, pricing controls, and packaging or warning requirements for THC content and serving size were absent.

**Figure 2.**
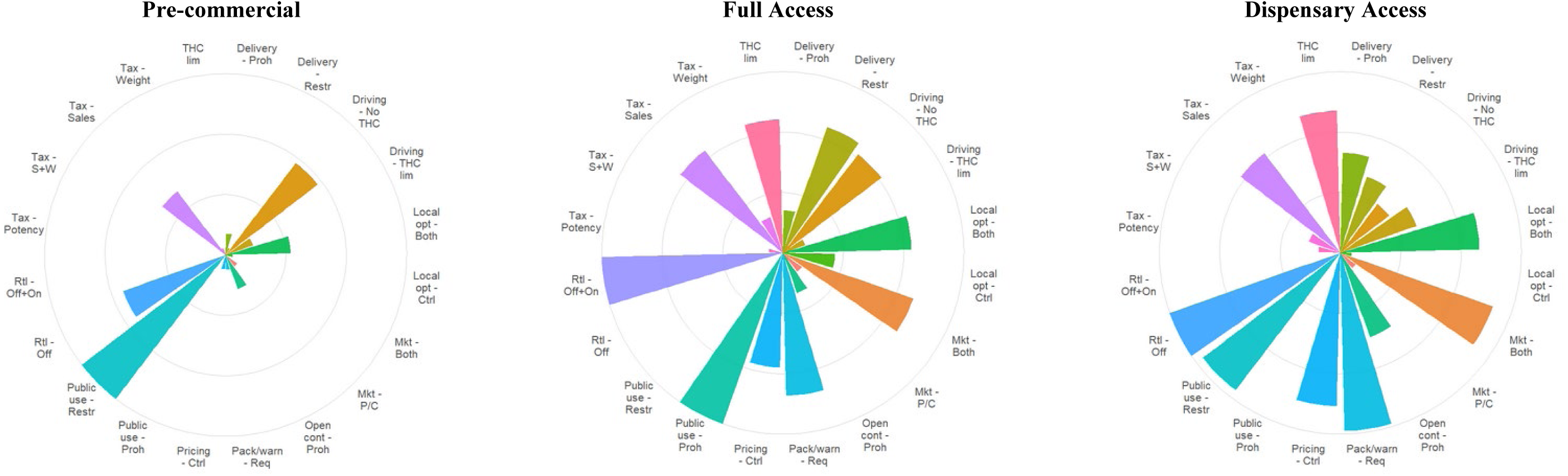
Endorsement probabilities of cannabis policy characteristics across latent classes Note: Each bar’s length is indicative of the probability of endorsement for a specific policy item, with the maximum possible endorsement probability being 1. Local opt - Ctrl: Local control authorized; Local opt - Both: Local opt-out and control authorized; THC lim: THC limits apply; Rtl - Off: Off-premises sales allowed only; Rtl - Off+On: On- and off-premises sales allowed; Public use - Restr: Public use restricted; Public use - Proh: Public use prohibited; Delivery - Restr: Home delivery allowed; Delivery - Proh: Home delivery prohibited; Tax - Sales: Sales-based taxes; Tax - Weight: Weight-based taxes; Tax - S+W: Sales + weight-based taxes; Tax - Potency: Potency-based taxes; Pricing - Ctrl: Price controls in place; Pack/warn - Req: THC & serving size on label required; Mkt - P/C: Ad Placement or content restricted; Mkt - Both: Ad Placement and content restricted; Driving - None: General driving prohibitions; Driving - No THC: No THC limits, evidence required; Driving - THC lim: THC limits specified; Open cont - No law: No law on open containers; Open cont - Proh: Open containers prohibited

The Full Access class (e.g. Colorado 2020-2024 and Nevada 2022-2024). This class showed a higher probability than the Pre-commercial subgroup of allowing local authorities to opt out of the statewide cannabis program and adopt additional regulations within their borders (p=0.71 vs. 0.29). The subgroup of state-years in this group had a high probability of implementing both content and placement advertising restrictions (p=0.76) and THC and serving size packaging or warning labeling requirements (p=0.79). All state-years in the Full Access subgroup allowed retail recreational cannabis sales for on-premises and off-premises consumption and restricted (but did not prohibit) public use. This subgroup was also characterized by a high probability of allowing home delivery (p=0.74) but lacked regulation on open container regulations for recreational cannabis in non-commercial vehicles (p=0.76).

Similar to the Full Access subgroup, state-years in the Dispensary Access (e.g. Colorado 2014-2019, Oregon 2015-202, Washington 2014-2024) had a high probability of allowing local authorities to opt out of the state cannabis program and to regulate within their borders (p=0.7) and to impose potency limits (p=0.79). However, in contrast to the Full Access subgroup, the Dispensary Access subgroup allowed only off-premises consumption, had a higher probability of imposing pricing controls (p=0.85), and was more likely to prohibit home delivery (p=0.55) compared to the Full Access subgroup (p=0.63 and p=0.24, respectively), and prohibited public use (p=0.95). In sensitivity analyses, the MLCA results for the 4-class solution were comparable to those from the standard LCA presented above (Figure S1). However, the standard latent class provided a more distinct separation of classes based on item endorsement probabilities, particularly for retail licensing types and cannabis use restrictions. In repeated analysis without DC (Figure S2 S3 in the Appendix), the LCA model for a 4-class solution also identified the subgroup of states without RCLs. However, the endorsement probabilities did not clearly separate subgroups of regulatory approaches. Therefore, we kept DC in the analysis, since it helped provide a clearer differentiation between subgroups and added substantive interpretation to the groups. Results were consistent with those presented in the main analysis when LCA models were restricted to states with recreational cannabis legalization (Figure S4), limited to the combined 2020–2024 period (Figure S5), and estimated separately for each year from 2020 to 2024 (Figure S6).

### 3.2 State characteristics with varying RCL approaches

Table 2 presents descriptive lagged characteristics of states (2019–2023) by the four RCL approaches in (2020–2024). The No RCL subgroup had the lowest past-year cannabis use prevalence (mean = 9.6%), compared to the three other subgroups, where past-year cannabis use prevalence ranged from = 14.2% to 16.1%. Cannabis sales per capita were the lowest in the No RCL subgroup and in the Pre-commercial subgroup. However, sales per capita were high in both the Full Access (mean = 177.2) and the Dispensary Access (mean = 138.4) subgroups. The Dispensary Access subgroup also had the longest time since MCL (mean = 20.5 years).

The proportion of state-years with at least one neighboring state with RCLs was highest in the Pre-commercial and Dispensary Access subgroups (68.8% and 71.4%, respectively). All RCL states had adopted Medicaid expansion, and nearly all had implemented comprehensive smoke-free air laws. The Dispensary Access subgroup had the highest proportion of state-years allowing ballot initiatives (69%) and the implementation of medical cannabis retail outlets (100%). Median household income was highest in the Pre-commercial and Full Access model subgroups compared to No RCL state-years. The percentage of votes for the Republican Party was higher in No RCL state-years (53.9%) than RCL state-years (32.2–43%). The Pre-commercial subgroup had the highest density of social organizations (22.7). Other demographic factors, such as racial composition, median age, and education, varied less across groups.

Results from the multinomial logistic regression model (Table 3) showed that past-month cannabis use among those aged 12 and older was associated with a greater likelihood of being in the Full Access subgroup versus No RCL (RRR = 1.78; 95%CI: 1.21, 2.62). The percentage of neighboring states with RCLs was not associated with any RCL subgroups. Greater experience with MCL, measured as the number of years since MCL, was associated with a higher likelihood of belonging to the Dispensary Access subgroup (RRR = 1.47, 95% CI: 1.02–2.12). Higher beer excise taxes were consistently associated with lower odds of being in any of the three subgroups with RCLs relative to the no RCL subgroup, suggesting that jurisdictions with more restrictive alcohol policies also tended to have more restrictive cannabis policies. A higher median age was associated with lower odds of being in the Full Access subgroup. Estimated associations for density of social organizations, urban population share, racial composition, household income, unemployment, and Republican vote share included the null across RCL subgroups.

**Table 3.** Associations between State-Year Characteristics (2019–2023) and Latent Classes of Cannabis Regulation, 2020–2024.

| Characteristic | Pre-commercial Model<br>RRR (95% CI) | Full Access<br>Model<br>RRR (95% CI) | Dispensary Access<br>Model<br>RRR (95% CI) |
| --- | --- | --- | --- |
| Past-month cannabis use (%) (Ages 12+) | 1.45* (0.99, 2.12) | 1.78*** (1.21, 2.62) | 1.27 (0.90, 1.78) |
| Annual cannabis sales per capita (\$) | 0.99 (0.97, 1.01) | 1.0 (1.002, 1.02) | 1.01 (1.001, 1.02) |
| Neighboring states with RCL (%) | 0.97 (0.93, 1.01) | 0.95 (0.88, 1.03) | 0.99 (0.95, 1.05) |
| Years since MCL | 1.03 (0.80, 1.33) | 1.25 (0.93, 1.67) | 1.47** (1.02, 2.12) |
| Beer excise tax (USD per gallon) | 0.93* (0.87, 1.01) | 0.93** (0.87, 0.99) | 0.94* (0.88, 1.01) |
| Votes in favor of the republican party in presidential elections (%) | 0.98 (0.88, 1.09) | 0.91 (0.79, 1.06) | 0.91 (0.76, 1.10) |
| Social organizations per 10,000 people | 1.04 (0.87, 1.25) | 0.66 (0.26, 1.66) | 0.68 (0.39, 1.20) |
| Urban population (%) | 0.94 (0.81, 1.09) | 1 (0.78, 1.28) | 0.99 (0.77, 1.27) |
| Median age (years) | 1.12 (0.65, 1.93) | 0.63** (0.39, 0.99) | 0.84 (0.55, 1.29) |
| Non-Hispanic white population (%) | 0.93 (0.85, 1.03) | 0.98 (0.87, 1.11) | 1.04 (0.90, 1.20) |
| Median household income (in \$1,000) | 1.07 (0.93, 1.22) | 1.06 (0.93, 1.22) | 1.02 (0.90, 1.16) |
| Unemployment rate (%) | 1.11 (0.78, 1.58) | 1.03 (0.85, 1.25) | 1.01 (0.86, 1.19) |

N=255
Reference group: No RCL group
RRR = Relative Risk Ratio; CI = Confidence Interval.
\*p < .10, \*\* p < .05, \*\*\* p < .01

## 4. DISCUSSION

The primary objective of this study was to characterize US state-years according to the RCL provisions in effect from 2013 to 2024. Using LCA, we identified four subgroups of state-years. Providing evidence of heterogeneity in state regulatory approaches: no RCL, Pre-commercial, Full Access, and Dispensary Access. This approach moves beyond a simple legalized versus non-legalized distinction for RCLs by offering a multidimensional classification system that differentiates states based on clusters of regulatory provisions.

The No RCL subgroup corresponded to state-years without recreational cannabis legalization and had zero probability of endorsing any provision included in the analyses. The Pre-commercial subgroup adopted a minimally regulated approach to commercial infrastructure, lacking a comprehensive commercial or public health framework, yet it had a high probability of prohibiting public use. This class was aligned with an early-stage form of legalization or with Washington, DC’s home-cultivation model.

Among the commercial RCL models, two distinct approaches emerged, refining the broad “commercial model” of RCLs used in prior studies (60). The Full Access subgroup corresponded to a regulated commercial model with greater leniency in where cannabis could be consumed—specifically, by permitting retail outlets with on-site consumption, as opposed to only retail outlets that sell cannabis products for off-site consumption. This distinction is relevant for public health, as regulated on-site consumption may reduce secondhand smoke exposure in private residences and public settings (61,62), but raises concerns about impaired driving (63) and the normalization of consumption, which can influence youth risk perceptions (64).

Similarly, state-years in this subgroup were more likely to allow home delivery while restricting (but not prohibiting) public use. By allowing retail sales for both off-site and on-site consumption, the Full Access approach established a state-level baseline for the development of cannabis consumption lounges, as documented in Nevada (65) and Alaska (66). In contrast, the Dispensary Access subgroup aligned with a commercial dispensary model that imposed stricter access and consumption restrictions than the Pre-commercial- and Full Access subgroups, allowing only off-site retail sales consumption and prohibiting public use.

The classification of state-years into distinct classes illustrated that RCL approaches varied across states and over time. Several early legalizing states were more often observed initially in the Pre-commercial group and later in the Dispensary Access or Full Access models. In contrast, late adopters were more likely to enter legalization with either the Full Access or Dispensary Access approach, bypassing the Pre-commercial stage. Our findings were consistent with previous research demonstrating heterogeneity in cannabis policy design, and over time (5,6,18). We extended prior frameworks by implementing a state-centered LCA rather than variable-centered composite-score approaches (6,42) that captured the overall degree of restrictiveness or permissiveness of a state. Through the LCA approach, we were able to inductively identify specific policy configurations that composite scores often mask. By characterizing specific combinations of RCL provisions rather than relying on broad composite measures, future research can identify which RCL approaches are most effective at mitigating the harms of cannabis use.

By examining the 2020-2024 period, after many states had already implemented RCLs, the descriptive and correlational analyses highlighted factors associated with different regulatory models during the policy-maturation period rather than with the initial decision to adopt a specific type of RCL. Within this context, higher cannabis use was associated with increased odds of adopting the Full Access approach relative to the no-RCL approach. This was consistent with previous studies suggesting that higher market demand may coincide with more access-oriented regulatory approaches (14,23). In contrast, the Dispensary Access approach was associated with a longer history of medical cannabis legalization and aligned with incremental policy-learning frameworks in which RCLs expand from existing medical systems but adopt tighter controls on consumption (5).

Contrary to our expectations from policy diffusion theory (19), the proportion of neighboring states with RCL was not significantly associated with adopting any RCL approach compared with the no-RCL subgroup. This result suggested that, during this period of widespread RCL adoption, the influence of neighboring legal markets on legalization decisions may have diminished or plateaued. Broader alcohol policy contexts appeared relevant: higher beer excise taxes were associated with lower odds of adopting any RCL approach, indicating that states with stricter alcohol control may also be more cautious towards cannabis liberalization.

Although the associations presented are not causal, these findings suggested that RCL approaches between 2020 and 2024 may have followed two different pathways: the Full Access approach may be primarily responding to demand, whereas the Dispensary Access approach may be more closely related to institutional experience, particularly a longer history of MCL,.

This study should be considered in light of several limitations. First, because of the large number of provisions and the limited sample size (51 jurisdictions), we were not able to conduct a latent transition analysis, which would have been the preferred approach for capturing transition probabilities across classes and over time. However, an LTA approach would have increased the number of parameters needed to estimate and reduced our ability to incorporate a wide list of RCL provisions. As the number of states implementing RCLs increases, future research should consider implementing an LTA. Second, our study is limited to state-level laws. Although our model included local control provisions, we were unable to account for heterogeneity within states (i.e., county or municipal level), which may lead to variation within our identified classes. Future research should expand on this work and incorporate county-level cannabis laws. Lastly, our study is limited to the provisions measured in the APIS database, using the most recent available data. However, these measures may mask meaningful variation within provisions. For instance, while we included whether the state imposed restrictions on advertising placement and content, we were unable to capture differences in the stringency or specificity of such restrictions because such details are not consistently available across states or years.

US states continue to debate and modify their cannabis regulatory structures, such as New York’s recent repeal of potency-based taxes on cannabis products (67). These ongoing changes highlight the importance of systematically characterizing how states structure their RCLs over time and assessing public-health implications of these shifts. Our exploratory findings suggested that two processes may be shaping policy approaches: one that appears largely market-driven and another influenced by institutional knowledge/policy learning.

Accordingly, the role of stakeholders (e.g., commercial industry lobbyists, public health officials, community advocacy groups, and consumers, among others) in policy development should also be evaluated. Although legalization efforts slowed in 2024, several states continue to consider legalization. The LCA approach in this study provided a foundation for identifying policy configurations that tend to co-occur in practice and for evaluating the effects of these policy clusters.

## 5. CONCLUSION

As legalization expanded nationally, RCLs have evolved into increasingly complex and differentiated frameworks across states. This study provided a multidimensional typology of how RCLs have evolved in the US. Unlike composite scoring systems that collapse multiple provisions into a single measure, the latent class approach used in this study enabled us to identify how recreational cannabis policy provisions cluster. The analyses revealed two distinct commercial subgroups that diverged on policies governing consumption restrictions and access, specifically the legality of on-site retail consumption, public use restrictions, and home delivery. Future research should build on this work and evaluate the impacts of distinct RCL approaches.

## Data Availability

All data produced in the present work are contained in the manuscript

