## Appendix for "Beyond legalization: characterizing distinct recreational cannabis regulatory approaches across US states, 2013-2024"

### Appendix S1: Data sources and operationalization

Table S1: Definitions and operationalizations of recreational cannabis law prohibitions in this study

| Provision | Description | Original values | Operationalization | Frequency | Source |
|---|---|---|---|---|---|
| *Governance* | | | | | |
| Regulatory agency | The name of the agency or agencies with authority to regulate recreational cannabis in that jurisdiction, as specified in the relevant statutes or regulations. | State-year specific, full list available at APIS | 0 Does not apply; | 481 | APIS |
| | | | 1 Not included; | 17 | |
| | | | 2 Regulatory agency includes public health or consumer protection agency | 4 | |
| Local authority | Authority conferred on localities to act with respect to recreational cannabis within their borders | Not available | 0 No or does not apply; | 510 | APIS |
| | | Local control | 1 Localities authorized to regulate recreational cannabis within their borders; | 16 | |
| | | Both | 2 Localities are authorized to opt out of the State-wide system or to regulate recreational cannabis within their borders | 86 | |
| Retail consumption | Retail consumption restrictions | Retail off premises: ‘nan ', No'<br>Retail on premises: ‘nan ', No' | 0 Does not apply or retail consumption on and off premises not allowed; | 494 | APIS |
| | | Retail off premises: 'Yes – License Required',<br>Retail on premises: 'No – License Required', | 1 Retail consumption off premises allowed, no on premises allowed; | 80 | |
| | | Retail off premises: 'Yes – License Required' | 2 Retail consumption on and off premises allowed | 38 | |
| Home delivery | Delivery, after sale, of recreational cannabis products to customers who are not physically present at a retail outlet is permitted or prohibited | nan<br>'No Law' | 0 Does not apply or no law; | 506 | APIS |
| | | 'Allowed with Restrictions'<br>'Allowed' | 1 Allowed with or without restrictions; | 58 | |
| | | 'Prohibited' | 2 Prohibited | 48 | |
| Public use | Consumption of recreational cannabis products in public (outside a private residence or other private location) is either prohibited or restricted | Nan; 'No Law' | 0 Does not apply or no law ; | 487 | APIS |
| | | ‘Restricted | 1 Restricted; | 38 | |
| | | 'Prohibited' | 2 Prohibited | 87 | |
| Home cultivation | Whether the jurisdiction permits or prohibits the delivery, after sale, of recreational cannabis products to customers who are not physically present at a retail outlet | Does not apply or no | 0 Does not apply or no | 495 | Martins et al. [1] |
| | | Allowed | 1 Yes | 117 | |
| Advertisement restrictions | Restrictions applicable to the advertising of recreational cannabis products for retail sale | ‘nan ', No' | 0 Does not apply or none | 511 | APIS |
| | | Content: Restrictions on what an advertising message says or conveys.<br>Placement: Restrictions on where or how an advertising message is conveyed. | 1 Content or placement | 14 | |
| | | Both (Content and Placement) | 2 Content and placement | 87 | |
| | | ‘nan ' | 0 Does not apply | 507 | APIS |

| | | | | | |
|---|---|---|---|---|---|
| Packaging requirements | Information about any requirements applicable to the physical packaging of recreational cannabis products offered for retail sale | 'Child-Proof/Resistant;<br>Tamper-Proof/Resistant' 'Other'<br>'Child-Proof/Resistant; Other' | 1 Serving size not required | 22 | |
| | | 'Child-Proof/Resistant; Serving Size'<br>'Child-Proof/Resistant; Serving Size; Other'<br>'Child-Proof/Resistant; Serving Size; Tamper-Proof/Resistant'<br>'Child-Proof/Resistant; Serving Size; Tamper-Evident'<br>'Child-Proof/Resistant; Serving Size; Tamper-Evident; Other'<br>'Child-Proof/Resistant; Serving Size; Tamper-Proof/Resistant; Other'<br>'Child-Proof/Resistant' 'Serving Size; Other' | 2 Serving size required | 83 | |
| Warning requirements | Information about any health and/or safety-related warning requirements that apply to the label or packaging of recreational cannabis products offered for retail sale | ‘nan ' | 0 Does not apply | 507 | APIS |
| | | 'Amount of THC; Breastfeeding; Impairment of Driving; Pregnancy; Presence of Cannabis or THC; Other'<br>'Amount of THC; Child Access; Impairment of Driving; Presence of Cannabis or THC; Other'<br>'Amount of THC; Presence of Cannabis or THC; Other'<br>'Amount of THC; Other'<br>'Amount of THC; Child Access; Impairment of Driving; Pregnancy; Presence of Cannabis or THC; Other'<br>'Amount of THC; Breastfeeding; Child Access; Impairment of Driving; Pregnancy; Presence of Cannabis or THC; Other'<br>'Presence of Cannabis or THC; Serving Size; Other' | 1 Amount of THC or serving size, but not together | 17 | |
| | | 'Amount of THC; Breastfeeding; Child Access; Impairment of Driving; Pregnancy; Serving Size; Other'<br>'Amount of THC; Child Access; Impairment of Driving; Presence of Cannabis or THC; Serving Size; Other'<br>'Amount of THC; Presence of Cannabis or THC; Serving Size'<br>'Amount of THC; Child Access; Impairment of Driving; Pregnancy; Presence of Cannabis or THC; Serving Size; Other'<br>'Amount of THC; Impairment of Driving; Presence of Cannabis or THC; Serving Size; Other'<br>'Amount of THC; Breastfeeding; Child Access; Pregnancy; Presence of Cannabis or THC; Serving Size; Other'<br>'Amount of THC; Presence of Cannabis or THC; Serving Size; Other' | 2 Amount of THC and serving size | 88 | |
| Pricing controls | The jurisdiction imposes one or more restrictions on the pricing practices of those engaged in selling recreational cannabis | | 0 Does not apply or none specified | 531 | APIS |
| | | Yes | 1 Yes | 81 | |
| THC limits | If the jurisdiction sets a potency limit specific to one or more product types | State-year specific included in a row or jurisdiction note, from APIS dataset. | 0 Does not apply or none specified | 533 | APIS |
| | | | 1 Yes | 79 | |
| State-wide retail outlet restrictions | | State-year specific included in a row or jurisdiction note, from APIS dataset. | 0 Does not apply or none specified | 565 | APIS |

| | | | | | |
|---|---|---|---|---|---|
| | Jurisdiction imposes limits on the number of retail licenses granted (per State or per individual) | | 1 Yes | 47 | |
| Taxation | Rate of tax imposed by the jurisdiction on recreational cannabis sales or transfers | nan | 0 Does not apply or no law | 497 | APIS |
| | | | 1 Sales based only | 83 | |
| | | | 2 Weight based only | 9 | |
| | | | 3 Combination of weight and sales | 12 | |
| | | '7% of gross receipts (producer-level); 10% for products with 35% THC level or less, 25% for products with more than a 35% THC level, and 20% for cannabis infused products (retail-level)'<br>'$0.00625 per mg of total THC for cannabis plant material; $0.0275 per mg of total THC for cannabis edible; $0.009 per mg of total THC for cannabis; 6.35% of sales (retail-level)'<br>'$0.005 per mg of total THC for cannabis flower material; $0.008 per mg of total THC for concentrates; $0.03 per mg of total THC for cannabis edible; 9% of sales (retail-level)' | 4 Potency based taxes | 11 | |
| Underage restrictions | Prohibitions specified by the jurisdiction with respect to cannabis and persons under 21 years of age | nan | 0 None or does not apply | 481 | APIS |
| | | 'Furnishing'<br>'Purchase; Furnishing'<br>'Purchase; Possession; Furnishing'<br>'Possession; Furnishing | 1 Consumption not explicitly prohibited | 18 | |
| | | 'Purchase; Possession; Consumption; Furnishing' | 2 Consumption explicitly prohibited | 113 | |
| Driving prohibitions | Whether the jurisdiction prohibits driving while under the influence of cannabis, together with any specified THC limit, as well as the evidentiary standard associated with that THC limit | nan | 0 Does not apply or none specified | 481 | APIS |
| | | 'Adult: No specific prohibition; Youth: No specific prohibition' | 1 General prohibitions apply | 29 | |
| | | 'Adult: Prohibited - THC limit and evidentiary standard not specified; Youth: No specific prohibition' | 2 No evidentiary limits specified | 64 | |
| | | 'Adult: Prohibited - 5 ng/mL limit (not per se) ; Youth: No specific prohibition' | 3 Limit specified (not per se) | 11 | |
| | | 'Adult: Prohibited - 2 ng/mL limit (per se) ; Youth: No specific prohibition'<br>'Adult: Prohibited - 5 ng/mL limit (per se) blood; 10\xa0ng/mL limit (per se) other bodily substance; Youth: No specific prohibition' | 4 Limit specified (per se) | 15 | |
| | | 'Adult: Prohibited - 5 ng/mL limit (per se)<br>Youth: Prohibited - 0 ng/mL limit (per se)' | 5 Limit specified (per se) & youth prohibitions | 12 | |
| Open container in the passenger compartment | Whether the jurisdiction permits or prohibits open containers of recreational cannabis or recreational cannabis products in the passenger compartments of non-commercial motor vehicles | Nan; | 0 Does not apply | 481 | APIS |
| | | 'No Law' | 1 No law | 85 | |
| | | 'Prohibited | 2 Prohibited | 46 | |

Table S2: Operationalization and data sources of state-level characteristics

| Characteristic | Measure | Data source |
|---|---|---|
| *Cannabis market* | | |
| Past-month prevalence of cannabis use (ages 12+) | Percentage of people 12 years or older who reported past-year marijuana use | NSDUH [2,3] |
| Cannabis sales per capita | Cannabis sales for recreational and medical cannabis sales by the population | State websites |
| Dispensaries per 10,000 | Number of state level dispensaries per 10,000 people | Weedmaps |
| *Cannabis policy adoption and diffusion* | | |
| Medical cannabis outlets operational | Indicator variable indicating whether medical dispensaries were operational | Martins et al. [1] |
| Neighboring state has RCL | Indicator variable whether a neighboring state had RCL | APIS |
| Years since MCL | Number of years since MCL | Martins et al. [1]- |
| *Substance use and health policy* | | |
| Beer excise tax | Excise tax rate levied on bee | APIS |
| Smoke free laws | Indicator variable whether the state implemented smoke-free laws | ANFR[4] |
| Medicaid expansion | Indicator variable whether the state implemented Medicaid Expansion | COEP |
| *Political characteristics* | | |
| Ballot initiatives allowed | Indicator whether the state allows for citizen initiative or referenda | NCLS[5] |
| Votes in favor of republican party in presidential elections (%) | Percent votes for republican party | MIT Election Data and Science lab [6] |
| Density of social organizations (per 10,000) | Social organizations (NAICS Code 813 corresponding to Religious, Grantmaking, Civic, Professional, and Similar Organization) per 10,000 people | Economic Census |
| State revenue as a share of expenses | Total expenditure as a share of total expenditure | Urban Institute |
| *Demographics* | | |
| Population non-Hispanic White (%) | Percent of the population non-Hispanic white | Census ACS |
| Population density (people per km2) | | Census ACS |
| Male (%) | Percentage of the population male | Census ACS |
| Population 18 to 34 (%) | Percent of the population aged 18 to 34* | Census ACS |
| Population 35 to 64 (%) | Percent of the population aged 35 to 64* | Census ACS |
| Population 65+ (%) | Percent of the population aged 65 or more* | Census ACS |
| *Socioeconomic characteristics* | | |
| Population 25 or older with High School+ (%) | Percentage of the population with high school diploma or more | Census ACS |
| Median household income (in $1,000) | Median household income | Census ACS |
| Unemployment rate (%) | Unemployment rate | U.S. Bureau of Labor Statistics[7] |

### Appendix S2: LCA implementation

Table S3: Model fit Summary for Standard LCA Model 1-5 Class Solution

| Classes | BIC | aBIC | CAIC | VLMR | *cmPk* |
|---|---|---|---|---|---|
| 1 | 7,635 | 7,565 | 7,657 | – | <.001 |
| 2 | 3,241 | 3,098 | 3,286 | **<.001** | <.001 |
| 3 | 3,040 | 2,824 | 3,108 | 0.17 | <.001 |
| 4 | **2,948** | 2,660 | **3,039** | **<.001** | **1** |
| 5 | 2,991 | **2,629** | 3,105 | 0.78 | <.001 |

Note: BIC = Bayesian Information Criterion; aBIC = sample size adjusted BIC; CAIC = consistent Akaike information criterion (lower values indicate better fit for all information criteria); VLMR = Vuong-Lo-Mendell-Rubin adjusted likelihood ratio test ($p<0.05$ indicates the k-class model fits significantly better than the k-1 model); cmPk = approximate correct model probability (values closer to 1 indicate higher classification certainty)

Table S4: Item endorsement probability by class for the 4-class solution

| Item | Description | No RCL | Pre-commercial | Full Access | Dispensary Access |
|---|---|---|---|---|---|
| Local opt - None | Neither local option nor local control are specifically authorized in the jurisdiction | 1 | 0.64 | 0 | 0.17 |
| Local opt - Ctrl | Local control is specifically authorized in the jurisdiction | 0 | 0.04 | 0.29 | 0.06 |
| Local opt - Both | Local option to opt-out and local control are specifically authorized in the jurisdiction | 0 | 0.32 | 0.71 | 0.77 |
| THC lim - No | No THC limits or does not apply | 1 | 1 | 0.26 | 0.22 |
| THC lim - Yes | Yes THC limits | 0 | 0 | 0.74 | 0.79 |
| Rtl - None | Does not apply or no consumption on premises/off premises allowed | 1 | 0.46 | 0 | 0 |
| Rtl - Off | Off premises consumption allowed, but not on premises consumption allowed | 0 | 0.54 | 0 | 1 |
| Rtl - Off+On | Off and on premises consumption allowed | 0 | 0 | 1 | 0 |
| Public use - None | Public use: Does not apply | 1 | 0.11 | 0 | 0.05 |
| Public use - Restr | Public use restricted | 0 | 0 | 1 | 0 |
| Public use - Proh | Public use prohibited | 0 | 0.89 | 0 | 0.95 |
| Delivery - None | Home delivery does not apply or no law | 1 | 0.86 | 0.03 | 0 |
| Delivery - Restr | Home delivery allowed with or without restrictions | 0 | 0.04 | 0.74 | 0.45 |
| Delivery - Proh | Home delivery prohibited | 0 | 0.11 | 0.24 | 0.55 |
| Tax - None | Does not apply or no law | 1 | 0.57 | 0 | 0 |
| Tax - Sales | Sales based only | 0 | 0.39 | 0.71 | 0.69 |
| Tax - Weight | Weight based only | 0 | 0.04 | 0.21 | 0 |
| Tax - S+W | Combination of weight and sales | 0 | 0 | 0 | 0.19 |
| Tax - Potency | Potency based taxes | 0 | 0 | 0.08 | 0.12 |

| | | | | | |
|---|---|---|---|---|---|
| Pricing - None | Does not apply or none | 1 | 0.93 | 0.37 | 0.15 |
| Pricing - Ctrl | Yes | 0 | 0.07 | 0.63 | 0.85 |
| Pack/warn - None | Does not apply | 1 | 0.93 | 0.21 | 0.02 |
| Pack/warn - Req | Amount of THC or serving size | 0 | 0.07 | 0.79 | 0.98 |
| Mkt - None | Does not apply or none | 1 | 0.93 | 0.11 | 0 |
| Mkt - P/C | Content or Placement | 0 | 0.07 | 0.13 | 0.11 |
| Mkt - Both | Content AND Placement | 0 | 0 | 0.76 | 0.89 |
| Driving - None | Does not apply | 1 | 0 | 0 | 0 |
| Driving – General | No specific prohibitions | 0 | 0.29 | 0.18 | 0.22 |
| Driving - No THC | No THC limits and evidentiary evidence specified | 0 | 0.57 | 0.68 | 0.34 |
| Driving - THC lim | THC limits specified | 0 | 0.14 | 0.13 | 0.45 |
| Open cont – None | Does not apply | 1 | 0 | 0 | 0 |
| Open cont – No Law | No law | 0 | 0.82 | 0.76 | 0.51 |
| Open cont - Proh | Prohibited | 0 | 0.18 | 0.24 | 0.49 |

### Appendix S3: Sensitivity Analyses

Figure S1: Results of the 4 latent class solution from the multilevel latent class model by state and year from 2013-2024

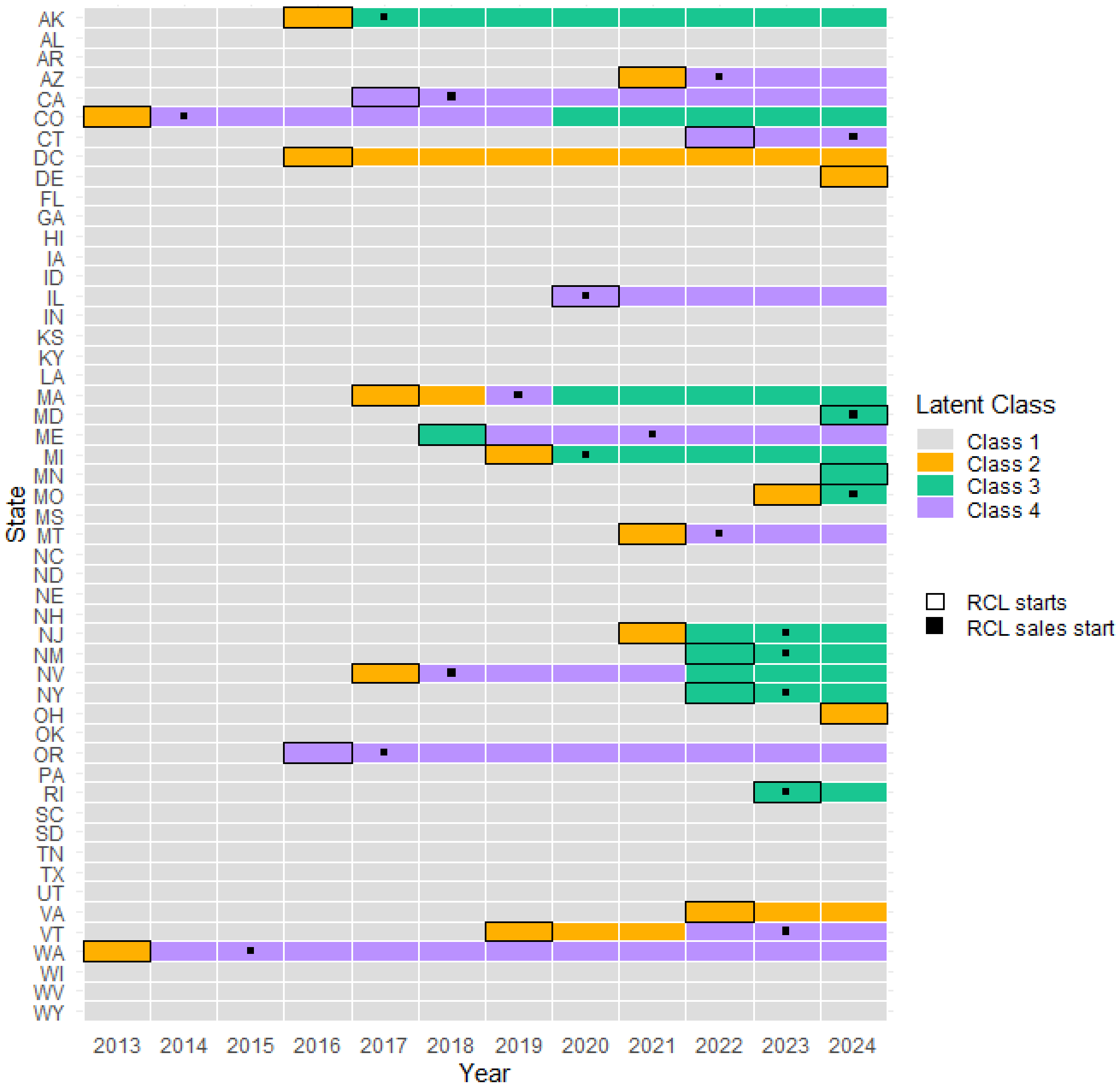

Figure S2: Results of the 4 latent class solution from the standard latent class model by state and year from 2013-2024, excluding DC

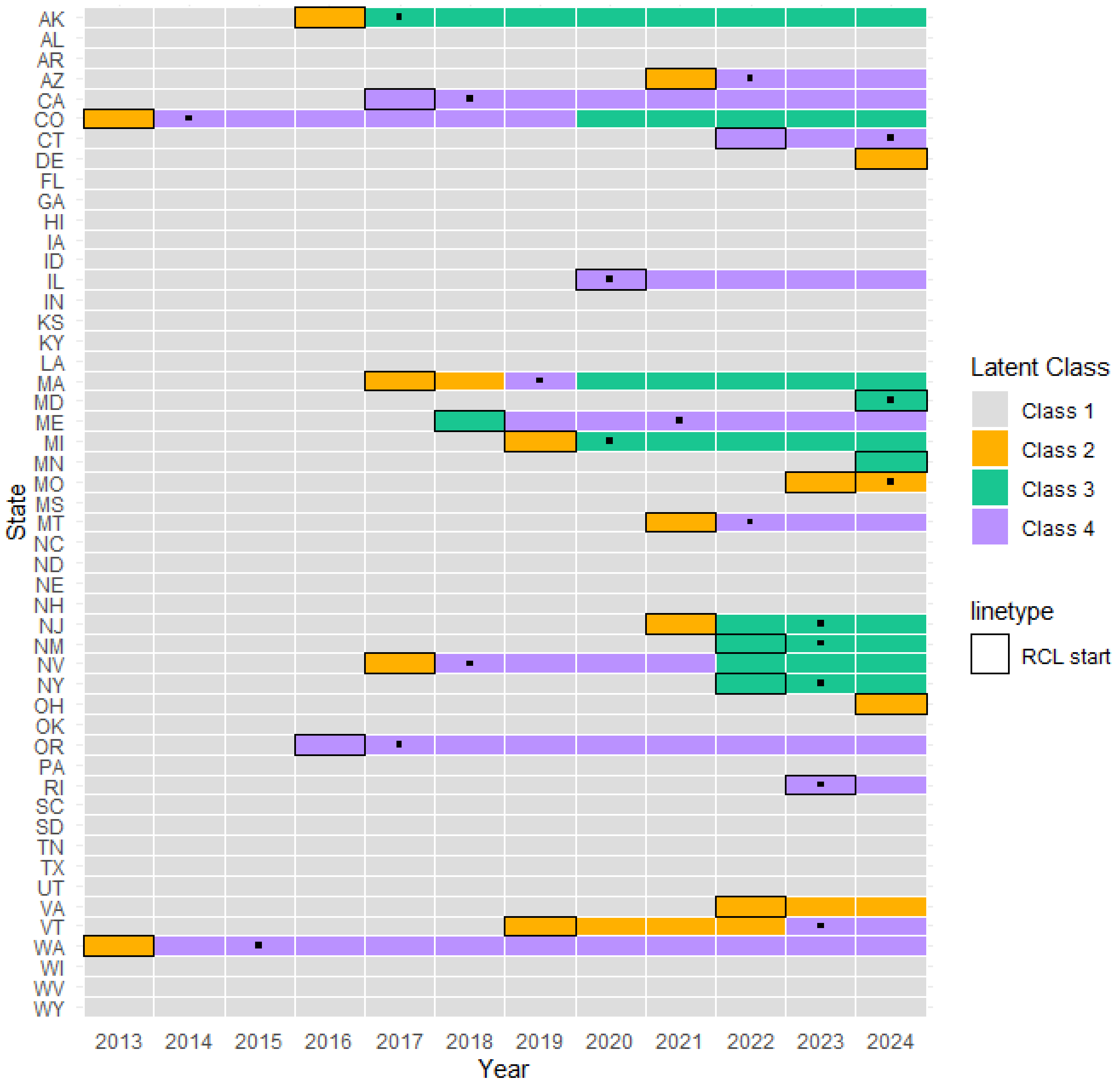

Figure S3: Results of the 4 latent class solution from the multilevel latent class model by state and year from 2013-2024, excluding DC.

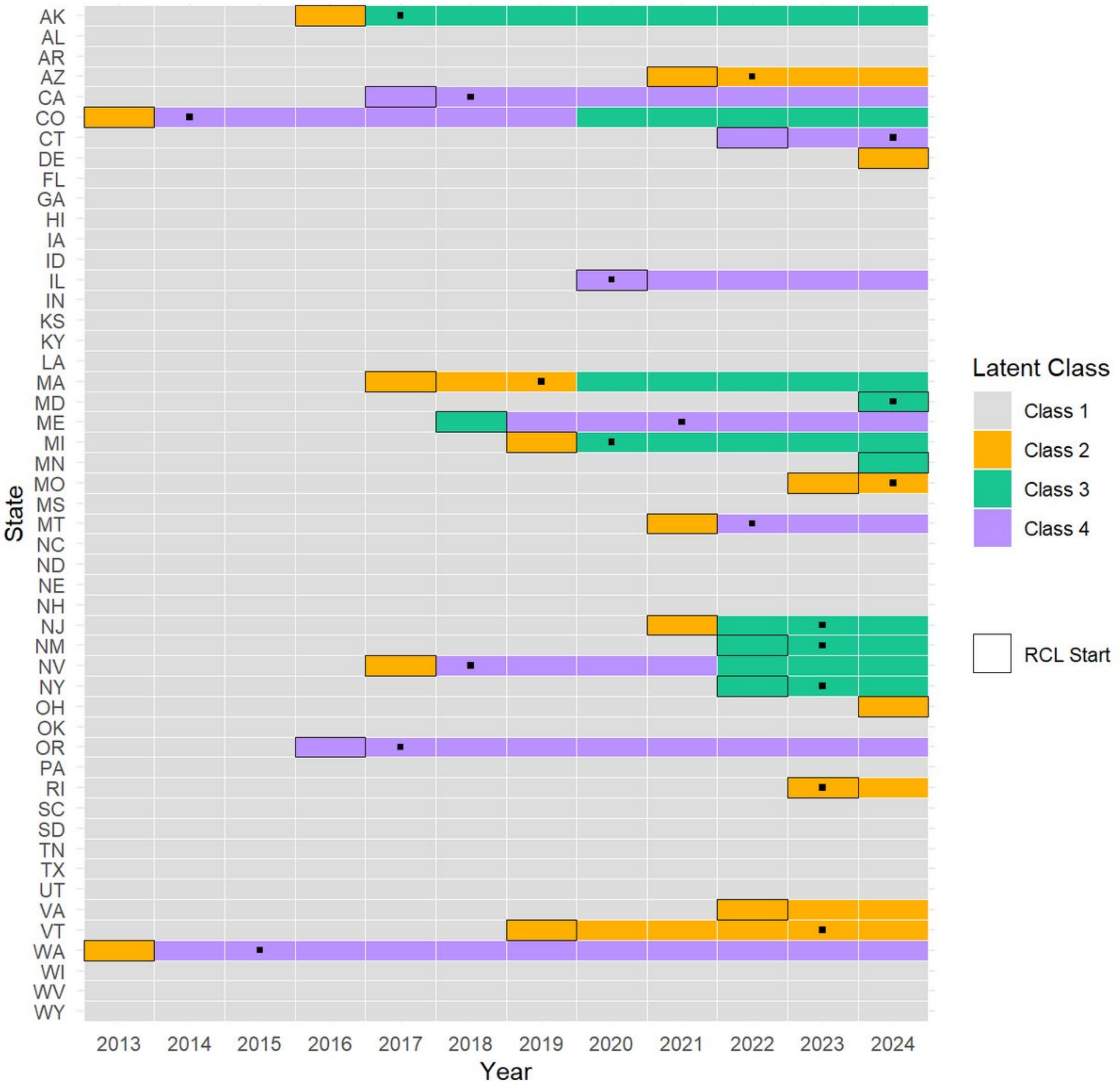

Figure S4: Latent Class by among states that legalized recreational cannabis use between 2012-2024

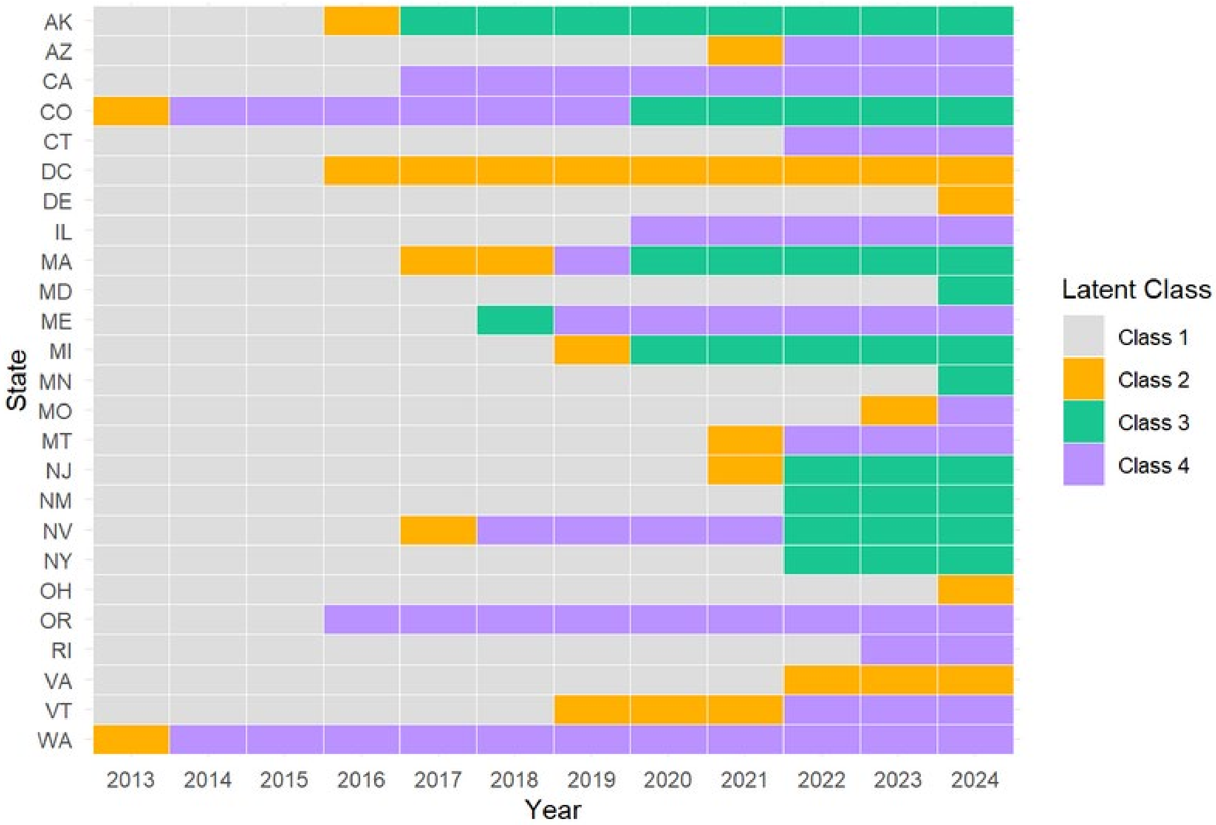

Figure S5: Results of the 4 latent class solution from the standard and multilevel latent class model by state and year from 2020-2024

**a) Standard LCA** **b) Multilevel LCA**

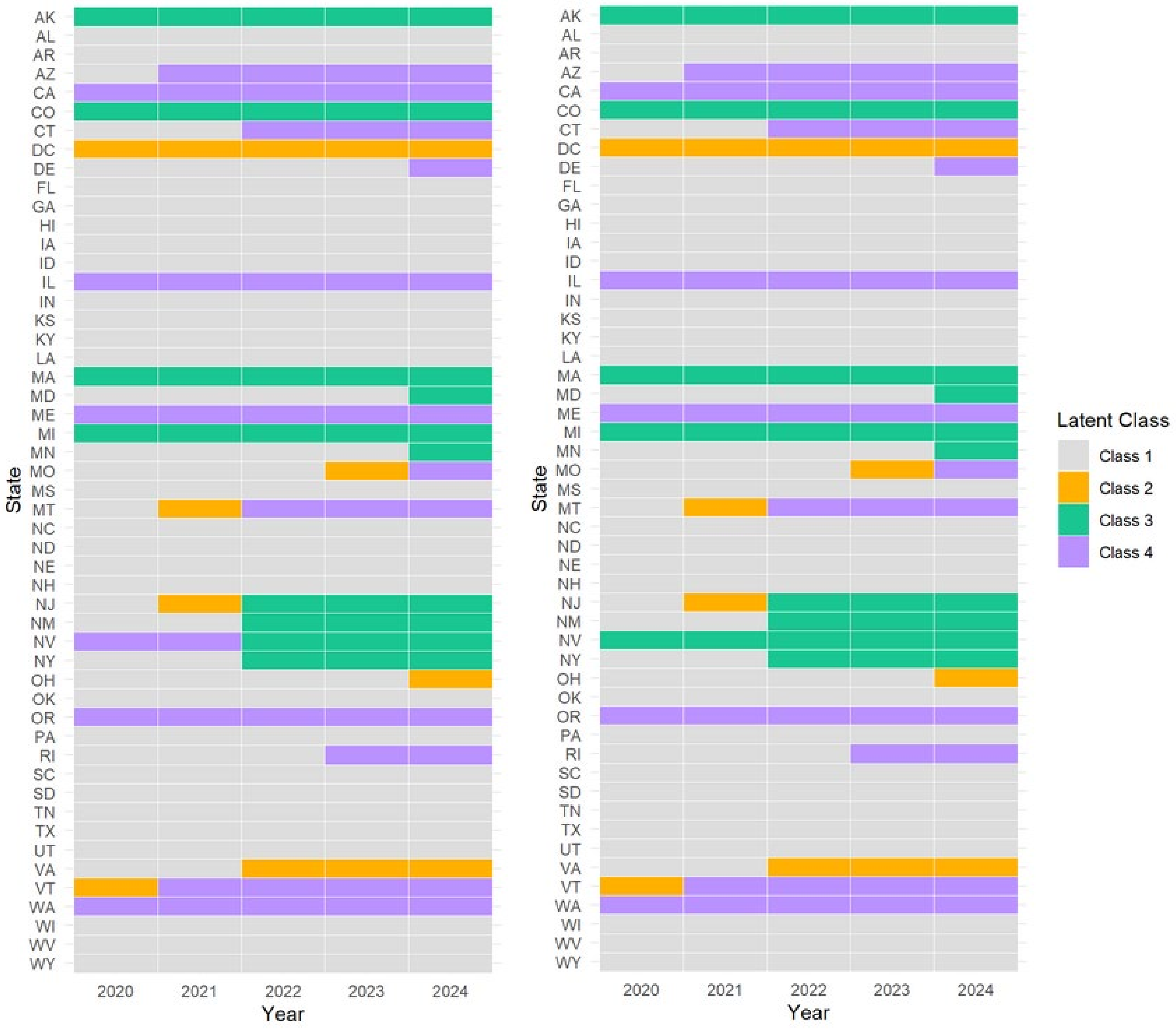

Figure S6: Results of the 4 latent class from 2020-2024, LCA conducted separately by year

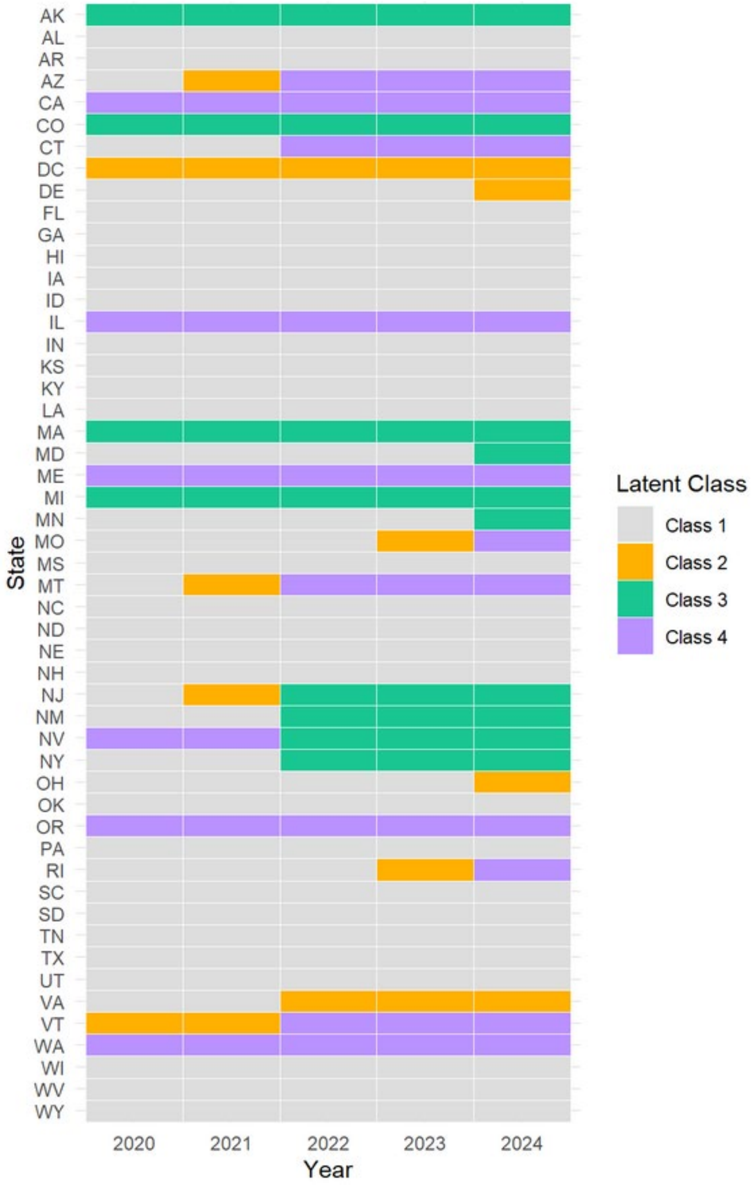
